# *epi*Liver a novel tumor specific, high throughput and cost-effective blood test for specific detection of liver cancer (HCC)

**DOI:** 10.1101/2021.02.07.21251315

**Authors:** David Cheishvili, Chifat Wong, Mohammad Mahbubul Karim, Mohammad Golam Kibria, Nusrat Jahan, Pappu Chandra Das, Md. Abul Khair Yousuf, Md. Atikul Islam, Dulal Chandra Das, Sheikh Mohammad Noor-E-Alam, Moshe Szyf, Wasif A. Khan, Mammun-Al-Mahtab

## Abstract

Robust cost effective and high-throughput tests for early detection of cancer in otherwise healthy people could potentially revolutionize public-health and the heavy personal and public burden of the morbidity and mortality from cancer. Several studies have delineated tumor specific DNA methylation profiles that could serve as biomarkers for early detection of Hepatocellular Carcinoma (HCC) as well as other cancers in liquid biopsies. Several published DNA methylation markers fail to distinguish HCC DNA from DNA from other tissues and other cancers that are potentially present in plasma. We describe a set of DNA methylation signatures in HCC that are “categorically” distinct from normal tissues and blood DNA methylation profiles. We develop a classifier combined of 4 CG sites that is sufficient to detect HCC in TCGA HCC data set at high accuracy. A single CG site at the *F12* gene is sufficient to differentiate HCC samples from thousands of other blood samples, normal tissues and 31 tumors in the TCGA and Gene Expression Omnibus (GEO) data repository (n=11,704). A “next generation sequencing”-targeted-multiplexed high-throughput assay was developed, which was used to examine in a clinical study plasma samples from HCC, chronic hepatitis B (CHB) patients and healthy controls (n=398). The sensitivity for HCC detection was 84.5% at a specificity of 95% and AUC of 0.94. Applying this assay for routine follow up of people who are at high risk for developing HCC could have a significant impact on reducing the morbidity and mortality from HCC.

## Introduction

Hepatocellular Carcinoma (HCC) is the fifth most common cancer world-wide [1] and is particularly prevalent in Asia; HCC occurrence is highest in areas where hepatitis B is prevalent which is a high-risk factor for HCC[2]. Follow up of high-risk populations such as chronic hepatitis patients and early diagnosis of transitions from chronic hepatitis to HCC would improve cure rates. The survival rate of hepatocellular carcinoma is currently extremely low because it is almost always diagnosed at the late stages. Liver cancer could be effectively treated with cure rates of >80% if diagnosed early^1^. Advances in imaging have improved noninvasive detection of HCC[3, 4]. However, current diagnostic methods, which include imaging and immunoassays with single proteins such as alpha-fetoprotein often fail to diagnose HCC early because of low accuracy and many early cancers are missed [2]. These challenges are not limited to HCC but common to other cancers as well.

Early detection of cancer in otherwise healthy patients requires noninvasive methods that could be administered to the wide public, are high throughput, don’t require sophisticated personnel or equipment to administer and are cost effective. Blood is an accessible biological sample and could be drawn in almost any location and shipped to centralized labs for further processing.

Molecular diagnosis of cancer is focused on tumors and biomaterial originating in tumor including tumor DNA in plasma [5, 6], circulating tumor cells [7] and the tumor-host microenvironment [8, 9]. Each of these approaches has its technical limitations. Detection of DNA from tumor origins in cell-free DNA (cfDNA) in plasma is an attractive approach, however it has its challenges, mainly the small and variable amount of circulating cell free tumor DNA as well as a requirement for an adequate method to differentiate tumor DNA from cfDNA that originated in other nonmalignant tissue in the plasma. Early studies focused on detecting tumor specific mutations by deep sequencing, however the variable abundance of specific mutations in tumors resulting from inter- and intra-tumor heterogeneity, which is further limited in the heterogenous population of cfDNA, reduces the sensitivity of such detection methods[10]. An alternative approach is examining unique tumor DNA methylation profiles by methylation specific PCR or next generation sequencing[11]. Aberrant DNA methylation profiles are a hallmark of cancer and a wide body of data has established that tumor DNA methylation profiles are dramatically different than their normal counterparts[12]. Early markers were based on a candidate gene approach utilizing candidate genes hypermethylated in cancer and were based on a limited set of tumor DNA methylation profiles. One of the most successful biomarkers that emerged from this approach is *Sept9* which is used in clinical practice for screening colorectal cancer[13].

Hypomethylation of HCC DNA is detectable in patients’ blood[14] and genome wide bisulfite sequencing was recently applied to detect hypomethylated DNA in plasma from HCC patients [15]. Methylscape is an assay that takes advantage of the global differences in the genomic distribution of methylation positions between cancer and normal tissues, which affect DNA physicochemical properties. The assay utilizes the differential interaction between gold and methylated and unmethylated DNA to detect cancer[16]. Initial studies with small number of samples suggest that it can detect cancer at close to 0.9 accuracy. However, this tool doesn’t provide information on specific cancer origin and it is unclear whether the global amount of tumor of cfDNA in plasma will be sufficiently abundant at early stages to be detected by this assay which examines global properties of DNA methylation.

More recently several groups performed a comprehensive comparative analysis of genome wide DNA methylation profiles of cancer, adjacent tissue DNA and blood in tumors and cfDNA to identify tumor specific DNA methylation profiles and compare these in tumor biopsies and cfDNA. An immunoprecipitation method with anti-5-methylcytosine antibody which analyzed methylomes of cfDNA in cancer patients revealed thousands of differentially methylated regions in cfDNA [17]. The methylation differences in cfDNA corresponded to DNA methylation differences in the tumors suggesting that DNA methylation signatures in tumor biopsies could be used to identify potential cfDNA tumor markers. A classifier composed of 300 differentially methylated regions (DMR) delineated by machine learning training classified cancer blood samples with high accuracy, sensitivity and specificity and the performance was similar between early and late-stage cancer, suggesting that certain tumor specific methylation profiles emerge early in cancer and could potentially be used for early cancer detection [17].

Genome wide bisulfite sequencing is a relatively costly procedure and requires significant bioinformatics analysis which makes it unfeasible as a screening tool. The challenge is therefore to delineate a small number of CGs that could robustly differentiate tumor DNA from nontumor DNA and to develop a high throughput cost effective assay that will enable the screening of wide populations in diverse geographic areas. While this study provides robust proof of principle for the utility of DNA methylation markers in early detection, it still requires 300 DMRs, which makes it difficult to develop into a widely used biomarker in a public health setting. A more recent pan cancer study has short listed 100,000 regions as tumor and tissue specific classifiers and validated them in a large multicenter clinical study. Although the test covers many cancer types it is significantly complex, and its specificity was low for early-stage cancers. The study strongly supports the idea that methylation profiles are more sensitive classifiers of cancer in cfDNA than tumor specific mutations [18].

In a different approach Xu et al., first compared DNA methylation in hepatocellular carcinoma HCC with blood DNA methylation Illumina arrays using publicly available datasets and established a DNA methylation panel, which was differentially methylated in HCC. This study compared methylation profiles of HCC tumor DNA and normal blood leukocytes and showed that matched plasma ctDNA and tumor DNA are highly correlated. The number of probes was reduced to 10 and they constructed a diagnostic prediction model which had a sensitivity of 85.7% and specificity of 94.3% for HCC in the training data set of 715 HCC and 560 normal samples and a sensitivity of 83.3% and specificity of 90.5% in the validation data set of 383 HCC and 275 normal samples [19]. However, the probes selected were not tested against normal DNA from other tissues that is present in cfDNA or against other cancer types.

The main challenge with many current approaches is that that they have not considered cfDNA from other tissues that is found in blood at different levels. Contaminating DNA from another tissue that has a similar methylation profile to a cancer tissue could potentially lead to false positives. In addition, past approaches have quantitatively compared DNA methylation in normal and cancer tissues. This quantitative difference is diluted when tumor DNA is mixed with different and unknown amounts of DNA from other untransformed tissues, which can cause false negatives. These deficiencies in current methods necessitate a different approach.

To address these challenges, we used thousands of methylation profiles in the TCGA and GEO publicly available data repository publica data collections to delineate DNA methylation positions that are consistently unmethylated in all the samples in the interrogated tissues and blood DNA. We then used this shortlist of ubiquitously unmethylated sites to identify sites that were highly methylated in a training set of HCC DNA methylation arrays. These sites exhibit a categorical binary difference between HCC and other tissues including blood. We shortlisted 4 CGs positions that are sufficient to classify HCC in a mixture of normal tissues and blood cells, which we termed “HCC-detect”. We then validated the methylation score composed of these sites in a data set composed of DNA methylation profiles of more than 700 HCC samples. We used a training dataset to discover a single CG site that was sufficient to differentiate HCC from 31 different cancers and normal cell types, which we termed “HCC-spec”. This was validated on a dataset of more than 8000 cancer samples from the public domain. We termed the combination of “HCC-spec” and “HCC-detect” “epiLiver”. The “epiLiver” test classified HCC samples in the public databases with high accuracy. We developed a targeted multiplexed high-throughput next generation bisulfite sequencing epiLiver test and applied it to plasma samples from 398 individuals from Dhaka city in Bangladesh. This novel test classified HCC patients at 95% specificity and 84.5% sensitivity and detected 75% of early stage A patients. Our study demonstrates the feasibility of a high throughput DNA methylation assay that interrogates a small number of CG sites to classify patients with HCC using a small amount of blood. Translation of such an assay into a routine early detection tool for people who are at high risk for developing HCC could have a significant impact on reducing the morbidity and mortality from HCC in high-risk populations.

## Results

### Delineating ubiquitously methylation resistant CG sites in blood and normal tissues

Tumor cfDNA is mixed with a background of normal and blood cfDNA in plasma at different and unpredictable amounts[20]. HCC DNA has both hypomethylated and hypermethylated regions that differentiate it from healthy DNA[21]. We reasoned that an ideal cancer marker would be a CG position that is ubiquitously unmethylated in normal tissues and blood but is methylated exclusively in tumors being categorically different than any normal cfDNA in blood. DNA methylation profiles could be heterogeneous across individuals; we therefore examined whether we could identify CG positions in publicly available DNA methylation arrays that are uniformly unmethylated across all the individuals and across 17 different somatic tissues. We first generated a list of 47981 CG positions that were hypomethylated in every single individual (beta=<0.1 and median <0.02) in 234 individuals in 17 different somatic tissues using Illumina 450K array data in GSE42752; GSE52955; GSE53051; GSE60185; GSE63704; GSE65821; GSE69852; GSE69852; GSE85464 and GSE85566. We then generated a list of 68260 unmethylated CG positions in blood DNA in each of the 312 individuals in GSE40279. We overlapped the two lists to obtain a list of CGs that are unmethylated in every single individual in both blood and 17 somatic tissues. To increase the robustness of the list and to exclude sites with residual variation in methylation across individuals that are derived from sex or age differences, we overlapped this list with a list of 60379 of unmethylated CGs in blood DNA in all 656 individuals males and females aged from 19 to 101 years (GSE40279). This overlap resulted in a final list of 28,775 CGs which are unmethylated across all individuals in multiple tissues at different ages and both sexes. The list of ubiquitously unmethylated sites were highly enriched for CG islands (10xe^-814^, Hypergeometric test), Transcription Start sites TSS200(7.7xe^-317^),1^st^ exon (3xe^-68^), 5’UTR(3.8xe^-27^), and Phantom High CG density promoters (5.22 fold enrichment 5.6xe^-395^) but depleted for the north and south shores of CG islands (3xe^-28^ and 3xe^-20^), enhancers (a 4.47 fold depletion 6.8xe^-145^), 3’UTR (a 13 fold depletion 3.8xe^-36^) and low CG density Phantom promoters (a 2.67 fold depletion 6.3xe^-5^). Thus, our list includes a highly selective group of CGs located in CG rich promoters that are uniformly “methylation resistant” across tissues and individuals.

### Discovery of 4 CG sites that classify HCC samples from healthy blood and other tissues; “HCC detect” markers

We then tested whether any of these ubiquitously “methylation-resistant” CGs become methylated in cancer. We used a dataset of Illumina 450K DNA methylation profiles from 66 HCC samples from all stages (GSE54503) and 77 control non-HCC liver samples (fibrosis and cirrhosis) (GSE61258) to generate a list of sites that show the highest methylation differential between HCC and control liver, limiting our analysis to the 28K methylation resistant CGs that we shortlisted. Remarkably, many of these ubiquitously methylation resistant CGs were methylated in HCC samples. 286 CG positions were methylated more than 20% in at least 50% of the HCC samples. A list of the top 20 CG sites with average difference in DNA methylation between HCC samples and non-HCC liver of above 0.2 (heatmap Fig. 1A) was further reduced by penalized regression to 4 CG sites; cg02012576 an intergenic region associated with the Checkpoint With Forkhead And Ring Finger Domain (CHFR) gene, cg03768777 at the 1^st^ exon of the Vasohibin 2 (VASH2) gene, cg05739190 at the 1^st^ exon of the Cyclin-J gene (CCNJ) and cg24804544 at the body of the Glutamate Receptor, Ionotropic, Delta 2 (Grid2) Interacting Protein 1 gene (GRID2IP). A weighted polygenic methylation score for HCC was computed by multivariable linear regression equation based on methylation values for these 4 CG positions in the training data (Table S1). The polygenic score significantly differentiated HCC and control samples (Fig.1B). A Receiver Operating Characteristic Curve (ROC) analysis performed on the calculated polygenic scores for the HCC and control samples shows an area under the curve of 0.9910 (Fig. 1C). Our training cohort included HCC samples from all stages with the goal of broad detection of cancer notwithstanding stage. We termed the 4CG marker set, “HCC detect”.

**Figure 1.**
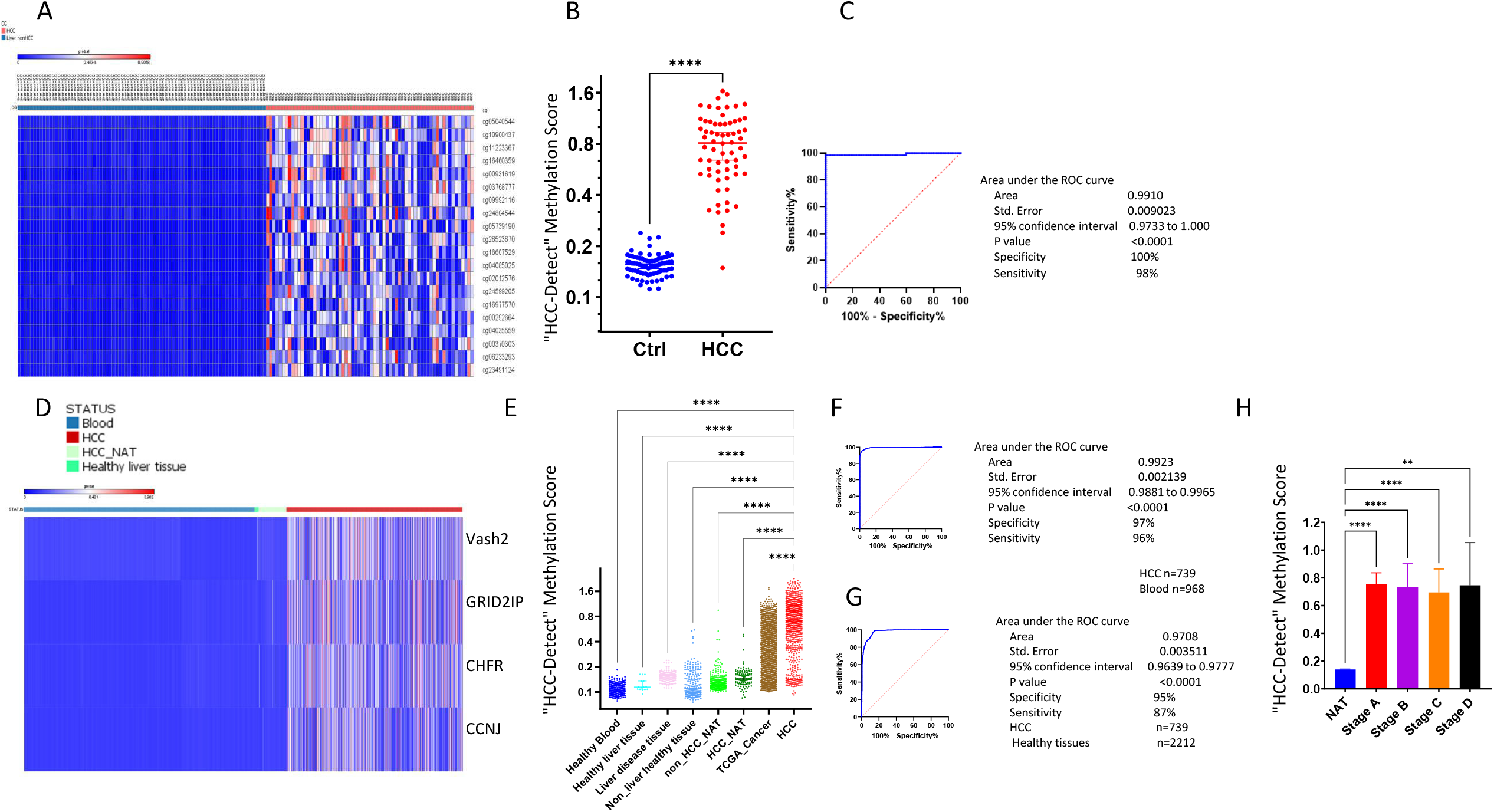
Training and validation of “HCC-detect” DNA methylation marker set. **A**. Heatmap showing methylation levels of top 20 CGs that are categorically different between noncancer liver samples (fibrosis) (n=79) and HCC samples (n=66) in the training cohort (GSE61258, GSE54503). **B**. Scatter plot of “HCC-detect” methylation scores calculated for HCC samples (n=66) and controls (n=79) in the training cohort (p<0.0001, Man-Whitney test, two tailed). **C**. ROC curve of “HCC-detect” methylation scores classifying blood and HCC samples from the training cohort. **D**. Heat map of methylation values for the 4 CG sites included in “HCC-detect” in the validation cohort of blood (n=968), heathy liver (n=16), liver NAT (n=116) and HCC samples (n=739) from TCGA and GEO data repository (see Table 1 for details). **E**. scattered plot of “HCC-detect” Methylation score (each spot represents one sample) in healthy blood (n=968), healthy liver tissue (n=16), liver disease (n=158), nonliver healthy tissues (n=234), NAT to nonHCC tumors (n=723), NAT to HCC (n=116), 31 cancers in TGCA (n=8754) and HCC (n=739). **F**. ROC curve of “HCC-detect” methylation scores classifying healthy blood (968) and HCC samples (739) in the validation cohort. **G**. ROC curve of “HCC-detect” Methylation scores of all healthy controls from different tissues and NATs (Table 1) (n=2212) and HCC (n=739) samples from the validation cohort. **H**. Median + confidence intervals of “HCC-detect” methylation scores in HCC NAT s (n=50) and different stages of HCC (Stage A 175, stage B 87, stage C 86, stage D 5) in the validation cohort (TCGA). (**** p<0.0001, ** p<0.001 Kruskal-Wallis nonparametric ANOVA with Dunn’s multiple comparisons test).

We validated “HCC detect” using DNA methylation 450K data for 793 HCC samples and 116 normal adjacent tissue (NAT) (heatmap Fig. 1D). We calculated the “HCC detect” score in 450K DNA methylation array data for healthy blood (n=968), healthy liver tissue (n=15), other liver disease (158), other healthy tissues (n=234), other normal adjacent tissues (721), cancers from 31 other tissues (8753), total n=11704 (Fig. 1E) (Table 1 and Table S2). The “HCC detect” polygenic score significantly differentiates HCC from all other groups as determined by one-way ANOVA after correction for multiple comparisons (F=793, p<0.0001, DF 11696; p<0.0001 for all comparisons) (Fig. 1E). AUC of 0.99 is computed by ROC curve analysis when HCC samples (n=739) are compared to Healthy blood (n=968) (sensitivity of 97% and specificity of 96%) (Fig. 1F), AUC of 0.97 when HCC (n=739) is compared to all healthy and NAT tissues including liver (n=2212) (specificity of 95% and sensitivity of 87%) (Fig. 1G), AUC of 0.95 when HCC samples are compared to 234 DNA methylation samples from healthy tissues (specificity of 95% and sensitivity of 85%), AUC of 0.92 when HCC (n=739) is compared to NAT of HCC (n=116) (specificity of 94% and sensitivity of 95%), AUC of 0.966 when HCC is compared to healthy liver tissue (specificity of 100% and sensitivity of 88%) and AUC of 0.87 when HCC is compared to 8753 samples from 31 different types of cancer (Table S2) (specificity of 90% and sensitivity of 64%). The HCC-detect methylation score detects early-stage HCC samples as well as late-stage HCC (Fig. 1H). These results validate that “HCC-detect” differentiates HCC samples at all stages from healthy tissues. Similar AUC values were obtained when equal weight was given to each CG in the detect score assuming that methylation at any of the 4 CGs is sufficient to classify a sample as HCC, though certain CGs are methylated (>20%) in a higher fraction of HCC samples than others (59% for Vash2, 57% for CHFR, 50% for GRID2IP and 44% for CCNJ) (Table S4).

**Table 1.** Data sets used for analysis.

| **Data Set** | **DNA origin** | **Description** | **Platform** | **N** | **Public Availability** |
| --- | --- | --- | --- | --- | --- |
| 1 | Non liver tissue | Healthy adjacent tissue | 450K | 723 | TCGA |
| 2 | Liver | Healthy liver tissue | 450K | 6 | GSE69852 |
| 3 | Liver | Healthy liver tissue | 450K | 10 | GSE76269 |
| 4 | Liver | Liver Cancer | 450K | 380 | TCGA |
| 5 | Liver | Healthy adjacent tissue | 450K | 50 | TCGA |
| 6 | Blood | Healthy | 450K | 656 | GSE40279 |
| 7 | Blood | Healthy | 450K | 312 | GSE61496 |
| 8 | Liver | nonHCC-Diseases | 450K | 158 | GSE61258 |
| 9 | Liver | Liver Cancer | 450K | 227 | GSE76269 |
| 10 | Liver | Liver Cancer | 450K | 66 | GSE75041 |
| 11 | Liver | Liver Cancer | 450K | 66 | GSE54503 |
| 12 | Non liver cancer tissues | 31 types of non HCC Cancers | 450K | 8754 | TCGA |
| 13 | Blood | cfDNA | MiSeq | 402 | This study |
| 14 | Lung | Normal | 450K | 26 | GSE63704 |
| 15 | Ovaries | Normal | 450K | 6 | GSE65821 |
| 16 | Stomach | Normal | 450K | 19 | GSE85464 |
| 17 | airway_epithelial_cells | Normal | 450K | 115 | GSE85566 |
| 18 | Breast tissue | Mamoplastic reduction | 450K | 15 | GSE60185 |
| 19 | Pancreas | Normal | 450K | 12 | GSE53051 |
| 20 | Non liver no cancer tissue | Kidney, prostate, bladder | 450K | 16 | GSE52955 |
| 21 | Colon | Normal | 450K | 19 | GSE42752 |

### Discovery of a single CG site whose methylation state classifies HCC samples correctly from tumors of different cell-type origins; “HCC-spec” marker

Several of the previously published early cancer detection DNA methylation biomarkers were were not tested across different cancers. Thus, these markers might detect several different kinds of cancers as well as HCC. The “HCC-detect” score developed for HCC preferentially detects HCC amongst 31 cancers in TCGA (Fig. 2A), however it detects other cancers as well, reducing the specificity and sensitivity of differentiating HCC from other tumors (specificity 90% and sensitivity 66%). To discover a set of markers that distinguishes tumors originating in the liver from other tumors we trained a cohort of 240 randomly selected DNA methylation samples from TCGA representing 16 different cancers, 10 HCC samples and 10 healthy blood samples. In this case, we didn’t limit our search to the 28,775 methylation-resistant CGs, in order not to miss liver-specific methylated CGs.

**Figure 2.**
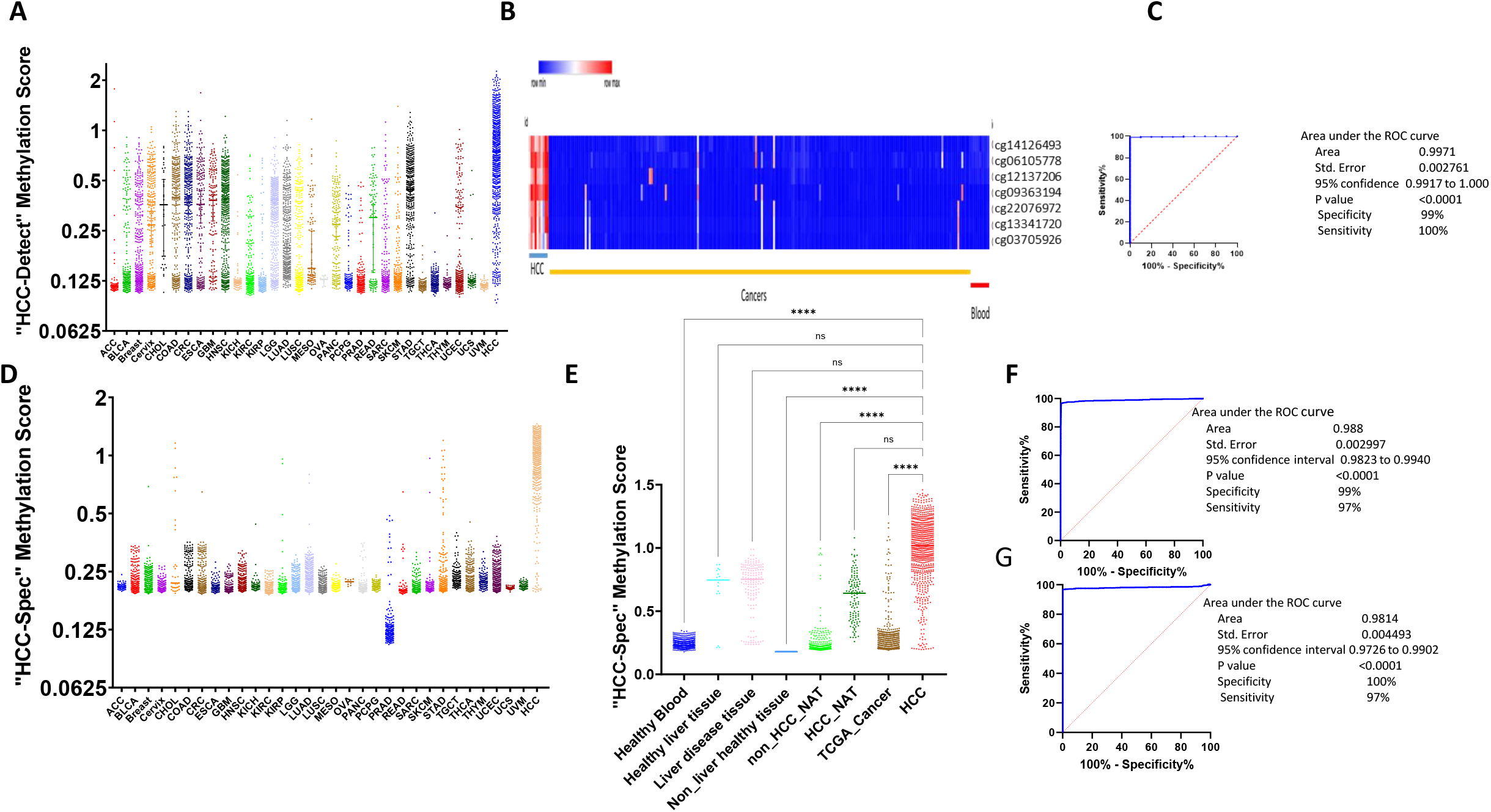
Training and validation of “HCC-spec” DNA methylation marker set. **A**. Scattered plot of “HCC-detect” methylation scores in 31 different cancer types and HCC (Table S2 for acronyms and number of samples). **B**. Heatmap of methylation levels of top 7 CGs shortlisted for discriminating HCC (n=10) from other cancers (10 randomized samples from each of 16 different cancers, ESCA, HNSC, TGCT, BLCA, Brain, Breast, CRC, PRAD, Stomach, Lung, Cervical, Pancreatic cancer, KIRC, KIRP, KICH, SKCM, and 10 blood) in the training cohort. **C**. ROC curve of “HCC-spec” methylation scores HCC (n=10) and other 16 kinds of cancer samples in the training cohort (n=210). **D**. Scattered plot of “HCC-Spec” methylation scores in 31 different cancer types and HCC (Table S2 for acronyms and number of samples per cancer). **E**. scattered plot of “HCC-spec” Methylation scores (each spot represents one sample) in healthy blood (n=968), healthy liver tissue (n=16), liver disease (n=158), nonliver healthy tissues (n=234), NAT to nonHCC tumors (n=723), NAT to HCC (n=116), 31 cancers in TGCA (n=8754) and HCC (n=739) (**** p<0.0001, n.s. nonsignificant, Kruskal-Wallis nonparametric ANOVA with Dunn’s multiple comparisons test). **F**. ROC curve of “HCC-spec” Methylation score classifying 31 other cancers (n=8754) and HCC samples (739) in the validation cohort. **G**. ROC curve of “HCC-spec” Methylation score classifying all blood samples (n=968) and HCC (n=739) samples in the validation cohort.

We calculated the differential methylation between the average methylation across the HCC samples and the average in all other cancers for each CG and its t-statistics. We shortlisted 7 CGs using a very strict threshold of (delta >0.5 and adjusted p value after Bonferroni correction for multiple testing of Q<10^−20^) (Heatmap in Fig. 2B). A multivariable linear regression with the 7 CGs as co-variates revealed that cg14126493 at the body of the *F12* gene has the largest effect. A weighted methylation score for *F12* computed by a linear regression equation (Table S3) classified all HCC samples correctly within a mixture of 240 samples in the training cohort. ROC curve (HCC:all other samples) revealed an AUC of 0.9973 with sensitivity of 99% and specificity of 100% (Fig. 2C). We designated cg1412693 as “HCC-spec” marker.

We then validated the “HCC-spec” as classifier of HCC DNA within a mixture of other tumors in a set of 11,692 samples that included 31 different cancers, non-malignant tissues and HCC samples (Table S2). “HCC-spec” is a liver specific marker. It differentiates HCC from 31 other cancers (scatter plot in Fig. 2D). The “HCC-spec” score is significantly different between HCC and healthy blood (p<0.0001), healthy tissues (p<0.0001), normal adjacent tissue (NAT) samples from 31 cancers (p<0.0001), and 31 cancers (p<0.0001) but is not significant between HCC and healthy liver tissue, liver-disease or HCC NAT (nonparametric Kruskal–Wallis one-way analysis of variance with Dunn’s multiple comparisons test) (Fig. 2E). The “HCC-spec” score accurately classified HCC samples (n=739) versus all other cancers (n=8754 for other cancers) with an AUC of 0.988 (99% specificity and 97% sensitivity) (Fig. 2 F). The AUC for HCC and normal blood (n=968) is similar 0.981 (100% specificity and 97% sensitivity) (Fig. 2G) and similarly the AUC for classifying HCC from healthy tissues (n=234) is 1 (100% specificity and 100% sensitivity). Remarkably, the DNA methylation level of a single CG site is sufficient to classify DNA as derived from liver tissue and not from other tissues or cancers. The “HCC spec” score however is not as accurate as expected for classifying HCC DNA and other non-malignant liver DNA as it is a liver-specific rather than cancer specific marker (Fig. 2E). The AUC for healthy liver tissue is 86% with specificity of 100% and sensitivity of 73%, and AUC for liver disease tissues of 0.84 with specificity of 95% and sensitivity of 71%. However, by combining the “HCC-detect” which accurately differentiates HCC and other liver DNA and “HCC spec” scores which differentiates HCC from other cancers (“HCC-detect”+”HCC-spec”=combined methylation score) we are able to accurately detect HCC DNA in a mixture of samples that included 31other cancers, normal tissue and liver tissues. AUC for HCC against all other tissues combined, including 31 cancers and liver tissues is 0.9862 with a specificity of 94% and sensitivity of 95% (Fig. 2G). At the threshold calculated by this ROC (a combined score of 0.87) the specificity against blood is 100%, against other 31 cancers is 95%, against normal tissue is 100% and against other cancers-NAT is 98.9%. However, at this threshold other liver tissues and liver disease DNA will be detected as well at the rate of 50%. To establish a threshold that differentiates HCC from other liver diseases we performed an ROC with HCC and other liver disease; the AUC is 0.937 at a sensitivity of 87% and specificity of 95% (using a higher threshold of a combined “HCC-detect” and “HCC-spec” score of 1.1).

We compared “HCC-spec” and “HCC-detect” DNA methylation markers (Fig. 3A) to two other extremely promising sets of HCC biomarkers that were recently described [22] [19] (Fig. 3B,C) using DNA methylation values for the respective CGs in Illumina 450K arrays from the 11701 samples described above. The heat maps presented in Fig. 3 show that although previously published markers display dramatic differences in methylation between HCC and HCC-NAT samples as previously reported, there is a high background of DNA methylation across other cancers and normal tissues. The combined “HCC-detect” and “HCC-spec” markers delineated here show a categorical differentiation between high methylation in HCC and extremely low methylation in other tissues and most cancers. While two of the “HCC-detect” markers are methylated to different extent in several cancers (Fig. 3B, C), *F12* is exquisitely methylated in HCC and liver disease samples but not in other cancers, normal tissues or blood (Fig. 3A).

**Fig. 3.**
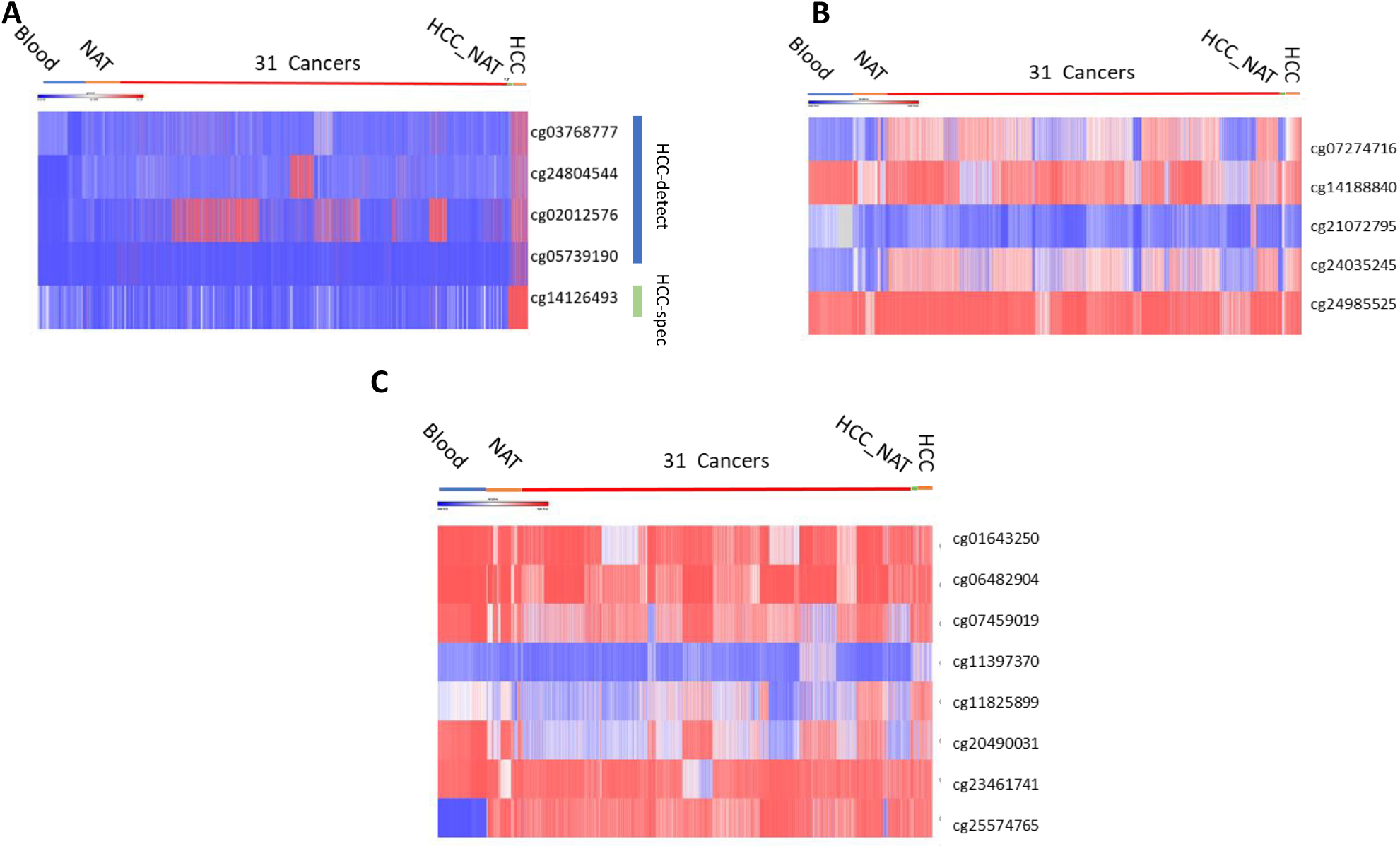
Comparisons of cancer type specificity of “HCC-detect and spec” combination and two previously published biomarkers. Heat map presentation of the methylation values for shortlisted HCC detection CG sites described here (**A**) in Liu et al[22] (**B)** and Zhang et al., **(C)** [19].

We tested whether the differential methylation of the 5 differentially methylated CGs discovered by analyzing DNA methylation data in Illumina 450K arrays will be differentially methylated in other publicly available methylation data derived by a different method. Wen et al., [23] examined both cfDNA and tumor tissue as well as NAT-HCC and plasma from liver cirrhosis and normal patients, by bisulfite conversion followed by methylated CpG tandem amplification and sequencing which enriches for methylated CGs (n=191) (GSE63775). We examined the count of methylated reads in genomic regions containing each of the 5 CGs of the “HCC-detect” and “HCC-spec” markers in this data set (57 tissue samples and 94 plasma samples). The 5 genes were significantly differentially methylated in all HCC tissue samples compared to HCC-NAT and in HCC plasma samples compared to plasma from cirrhosis and normal livers (Fig. 4A,B,D.E insets) with the exception of *GRID2IP* which showed a difference in methylation which didn’t reach significance in plasma because of the low number of reads in the serum sample (Fig. 4. C inset), but nevertheless it was significantly differentially methylated in HCC tissue samples (Fig. 4C). These data confirmed in an independent data set that the “HCC-detect” and “HCC-spec” CGs are methylated in HCC and in plasma cfDNA in HCC patients.

**Fig. 4.**
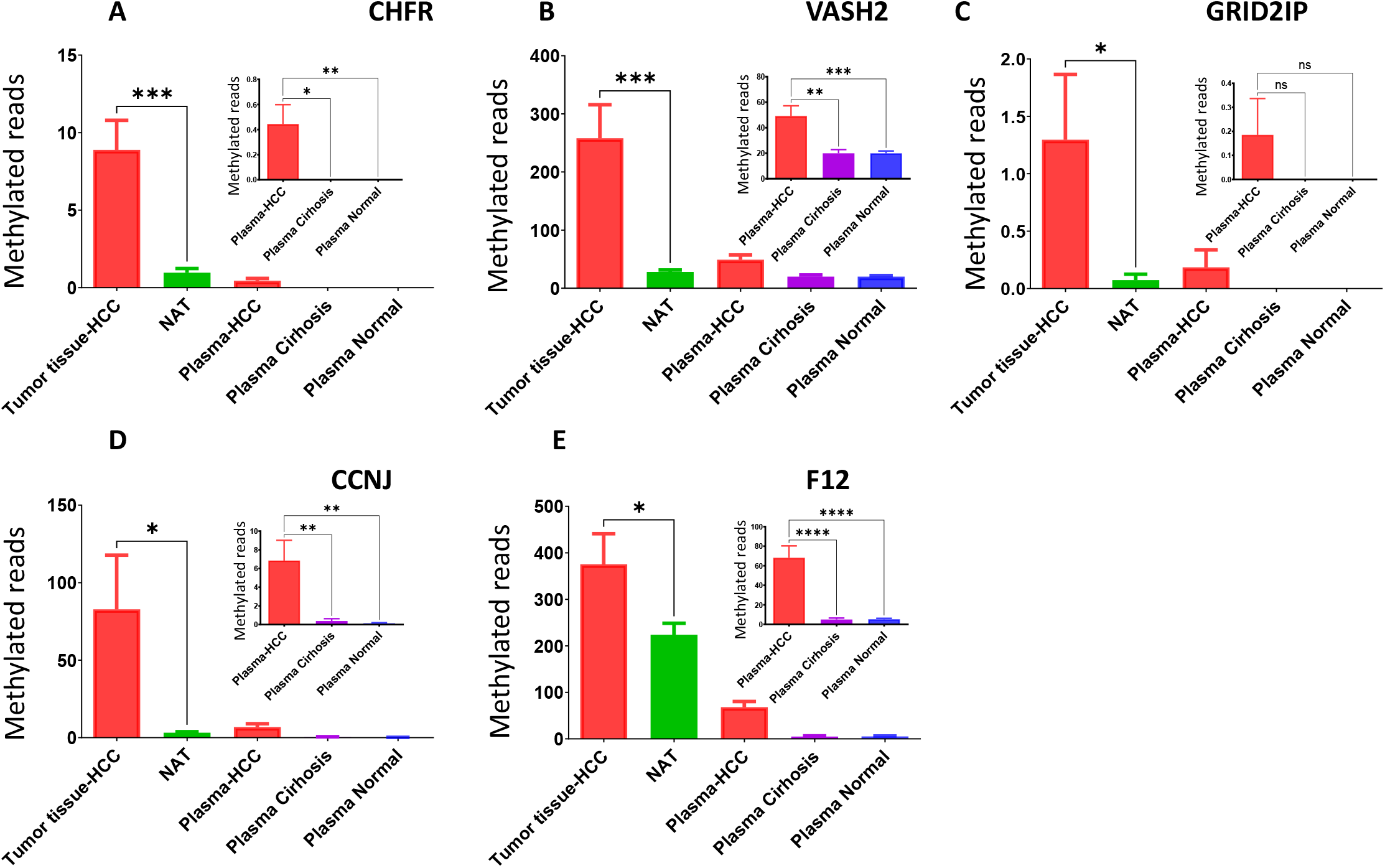
Validation of differential methylation of “HCC-spec” and “HCC-detect” CG sites in (GSE63775). Count of methylated reads in genomic regions containing each of the 5 CGs of the “HCC-detect” and “-spec” markers in this data set (57 tissue samples and 94 plasma samples). **A**. *CHFR* **B**. *VASH2*. **C**. *GRID2IP*. **D**. *CCNJ*. **E**. *F12*. Insets represent an enlargement of the charts for plasma cfDNA. For tumor tissues, significance was determined by nonparametric two tailed t test. For plasma, we performed Kruskal-Wallis nonparametric ANOVA with Dunn’s multiple comparisons test (**** p<0.0001, *** p<0.001, ** p<0.01, *p<0.05, n.s. nonsignificant).

### Validation of HCC-detect and HCC-spec DNA methylation in a clinical study examining plasma cfDNA in 398 people (Clinical trial gov ID:NCT03483922)

We recruited 402 people from Dhaka city in Bangladesh which included healthy controls, chronic hepatitis B patients as well as patients at different stages of HCC from stage 0 to stage D according to the EASL–EORTC Clinical Practice Guidelines (Clinical data summary Table 2). In difference from examining DNA methylation in a tumor biopsy where a significant fraction of DNA is derived from the tumor (as in the TGCA methylation database), tumor DNA in plasma is mixed with DNA from potential other sources and the extent of dilution of tumor DNA in other DNA is unknown. Thus, the level of methylation of plasma DNA might reflect an unpredictable dilution of tumor DNA;the level of methylation of cfDNA is therefore a function of the state of methylation of DNA in the tumor and the unknown and stochastic mixture with other DNA. Thus, it is anticipated that the level of methylation is lower than what we derived from examining tumor DNA methylation data. However, if the methylation profile of the tumor DNA is categorically distinct from the methylation profile of other potential sources of DNA in plasma as anticipated by the analyses above (Fig. 1-3), we expected that it would be detectable even on a high background. We used bisulfite mapping combined with next generation sequencing, which provides DNA methylation profiles at a single DNA molecule resolution.

**Table 2.** Demographics of study participants. Significant difference from the control group was tested by Kruskal-Wallis non-parametric one-way ANOVA and was adjusted by Dunn’s multiple comparisons test. The two HCC stage O samples were included for statistics in the HCC A group.

|  |  | **Age** |  | **Sex** |  | **Alcohol** |  | **Smoking** |  |
| --- | --- | --- | --- | --- | --- | --- | --- | --- | --- |
|  | N | mean | significance | male/female | significance | fraction users | significance | fraction users | significance |
| **Control** | 49 | 26.14 |  | 42/9 | 0.9999 | 0.2 |  | 0.32 |  |
| **CHB** | 51 | 28.71 | 0.9999 | 42/7 | 0.9999 | 0.21 | 0.999 | 0.35 | 0.999 |
| **HCC O** | 2 | 62 | NA | 2/0 | 0.9999 | NA | NA | NA | NA |
| **HCC A** | 32 | 49.15 | <0.0002 | 27/5 | 0.9999 | 0.088 | 0.353 | 0.64 | 0.0205 |
| **HCC B** | 86 | 49.12 | <0.0003 | 72/14 | 0.9999 | 0.069 | 0.0473 | 0.52 | 0.13999 |
| **HCC C** | 106 | 49.34 | <0.0004 | 85/21 | 0.9999 | 0.037 | 0.0047 | 0.54 | 0.0533 |
| **HCC D** | 77 | 52.45 | <0.0005 | 76/64 | 0.9999 | 0.064 | 0.0425 | 0.66 | 0.0012 |

We developed a multiplexed targeted amplification next generation sequencing bisulfite mapping assay, that measures the state of methylation of regions spanning 100 to 200 bp around the “HCC-spec” and “HCC-detect” CGs in up to 200 people in parallel. We first examined whether the 5 CG positions in the genes that we have selected are differentially methylated in plasma cfDNA derived from HCC patients in comparison to plasma from healthy people or from people with chronic hepatitis B (n=402, 4 samples with sequencing reads below 100 were removed from the analysis and we remained with 398 informative samples). The analysis included 46 healthy controls, 49 Chronic hepatitis, and 302 patients with HCC at different stages: Stage 0-2, Stage A-34, Stage B-86, Stage C-106 and Stage D-76). All “HCC-spec” and “HCC-detect” CGs were significantly more methylated in HCC patients than in healthy and chronic hepatitis B patients plasma (one way ANOVA with correction for multiple comparisons) with the exception of CCNJ which was significantly more methylated in HCC patients than in chronic hepatitis but was nominally significant in HCC to controls comparison; while median methylation in control and chronic hepatitis B heterogenous was slightly above 0%, methylation of up to 50 to 80 % was noted in the HCC samples (Fig 5A scattered plots). Hypermethylation was noted even at early HCC stages (Fig S1A) similar to the results obtained in the TCGA HCC dataset (Fig. 1H). These data confirmed that the “HC-detect” and “HCC-spec” CGs selected using TGCA tumor methylation data are differentially methylated in HCC patients’ plasma.

**Fig. 5.**
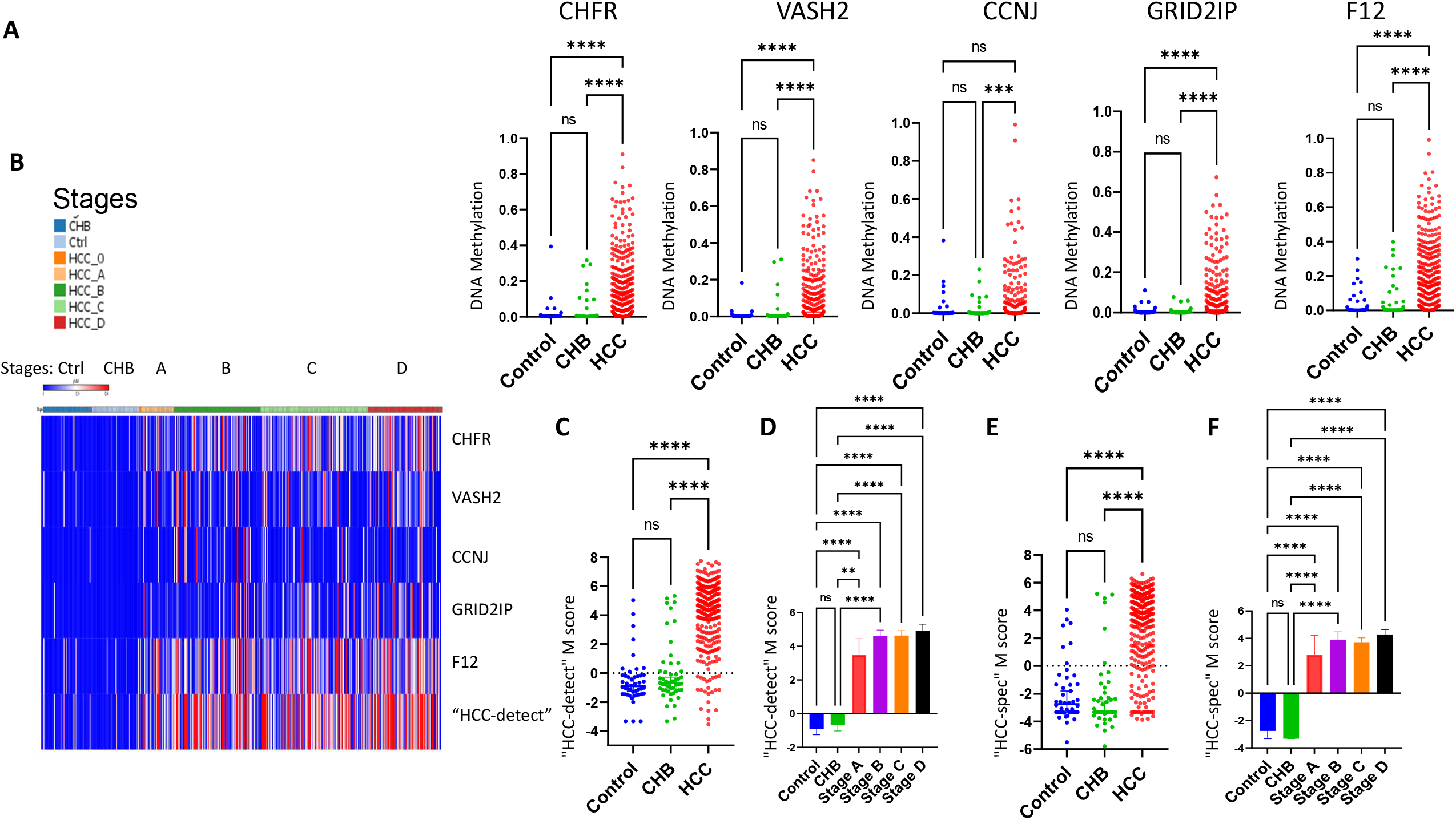
Validation of “HCC-detect” and “HCC-spec” in plasma samples in the Dhaka clinical study. **A**. Differential methylation of CGs included in the “HCC-detect” markers (cg02012576 (CHFR), cg03768777 (VASH2), cg05739190 (CCNJ), cg24804544 (GRID2IP) and “HCC-spec” cg14126493 (F12) in healthy controls (n=46), CHB (n=49), HCC (302). Beta values for each sample are presented in the scatter plot. **B**. Heatmap depicting the median methylation for each of the 5 regions amplified included in “HCC-detect” and “HCC-spec”. “HCC-detect” row shows the sum of median methylation values for each sample. The color codes for the groups are listed in the legend. **C**. Scattered plots of the “HCC-detect” M scores for control (n=46), CHB (n=49) and HCC (302). **D**. Median “HCC-detect” M scores for healthy controls (n=46), CHB (n=49) and 4 stages of HCC (Stage A+O-36, Stage B-86, Stage C-106 and Stage D-76)+/- confidence intervals. **E**. Scattered plots of the “HCC-spec” M scores for control, CHB and HCC. **F**. Median “HCC-spec” M scores for healthy controls, CHB and 4 stages of HCC +/- confidence intervals. Significance was determined by Kruskal-Wallis nonparametric one-way ANOVA with Dunn’s multiple comparisons test (**** p<0.0001, *** p<0.001, ** p<0.01, *p<0.05, n.s. nonsignificant).

Targeted sequencing allows capturing the methylation state of several CG in the proximity of the CGs that were selected using the TCGA data. We noted that in all 5 regions, hypermethylation was not limited to the 5 CGs selected in the 450K arrays and that there was a high correspondence in methylation levels of the CG included in the “HCC-detect” and “HCC-spec” sets and proximal CGs (heatmap Fig. S1B). To evaluate the consistent methylation state across the regions, we computed the median methylation in the amplified region for each of the 5 genes (heatmap Fig. 5B). We used median rather than average to exclude situations where a high average is driven by a spurious high methylation of a single CG. Median values of percentage methylation from 0 to 100 were normalized (log 2) and a “HCC-detect” M score was computed from the SUM of normalized medians of *CHFR, VASH2, CCNJ* and *GRID2IP* regions giving equal weight to each region. Similarly, the “HCC-spec” M score detecting HCC specificity versus other cancers was computed from the median methylation of the *F12* region. Both scores “HCC-detect” and “HCC-spec” significantly differentiated the HCC group from either the healthy control or chronic hepatitis B groups (one-way non-parametric ANOVA with correction for multiple comparisons) (Fig. 5C, 5E) but there was no significant difference between the chronic hepatitis B (CHB) and the healthy control groups. Importantly, both the “HCC-detect” M scores and the “HCC-spec” M scores were significantly different from control and chronic hepatitis B groups at early and late HCC stages (Fig. 5D, F): there are no significant difference between HCC stages (one-way non-parametric ANOVA with Dunnett correction for multiple comparisons).

To examine the biomarker quality of the” HCC-detect” M score we analyzed its receiver operator characteristics. The AUC for the “HCC-detect” score (302 HCC patients and 46 controls) was 0.93, the specificity 91% and sensitivity 89% (Fig. 6A).

**Fig. 6.**
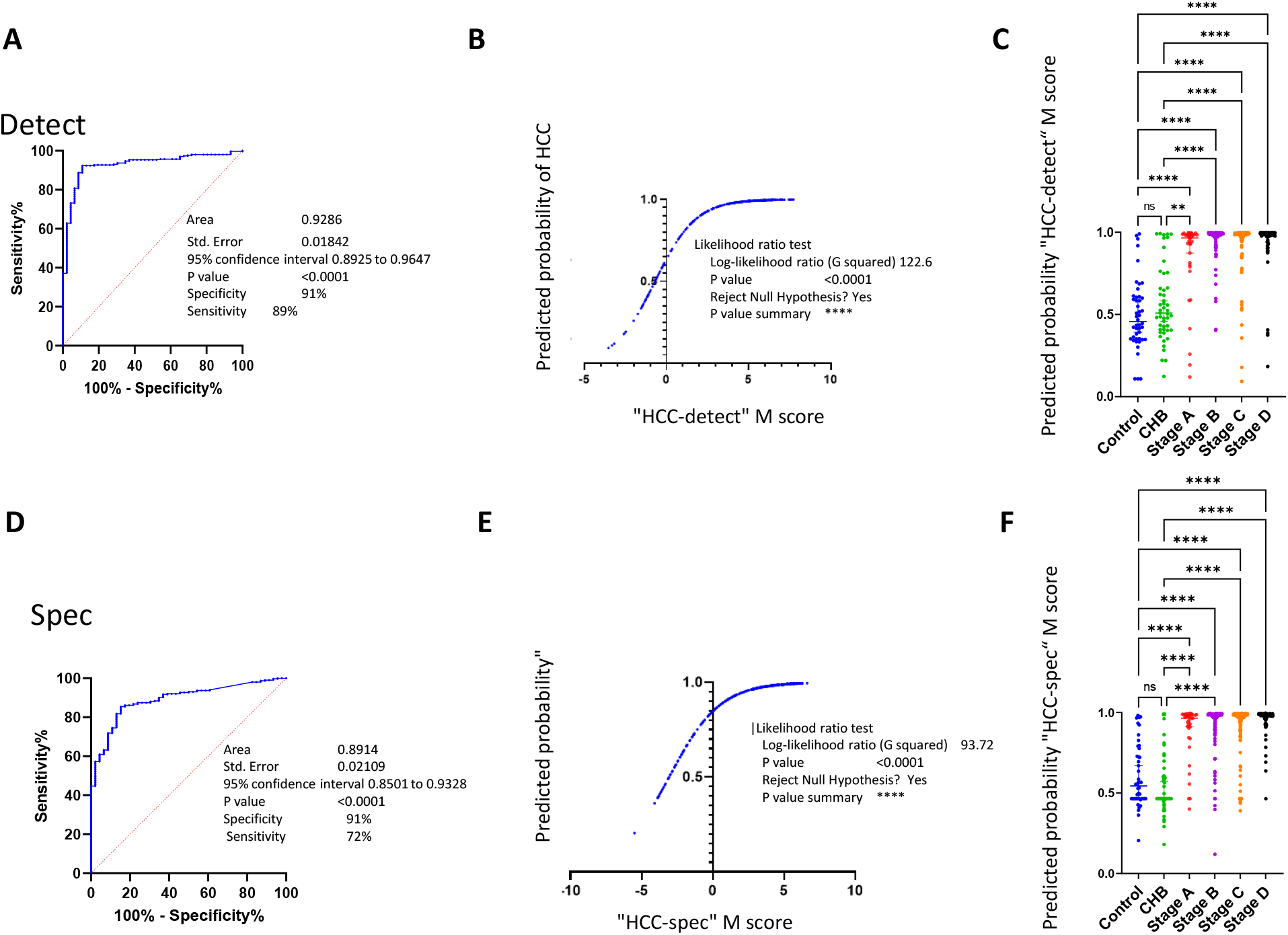
Biomarker characteristics of “HCC-detect” and “HCC-spec” M scores. **A**. ROC curve of “HCC-detect” M scores classifying HCC in healthy control (n=46) and HCC cases (n=302) in the Dhaka clinical study. **B**. Logistic regression curve plotting the predicted probability of HCC as a function of “HCC-detect” M score. **C**. Predicted probabilities (0 to 1) for each of the samples from the Dhaka clinical study calculated using the logistic regression equation for the “HCC-detect” M scores. **D**. ROC curve of “HCC-spec” M score classifying healthy control (n=46) and HCC cases (n=302) samples in the Dhaka clinical study. **E**. Logistic regression curve plotting the predicted probability of HCC as a function of “HCC-spec” M score. **F**. Predicted probabilities (0 to 1) for each of the samples from the Dhaka clinical study calculated using the logistic regression equation for the “HCC-spec” M score. The clinical study included (controls (n=46), CHB (n=49) and 4 stages of HCC (Stage A+O-36, Stage B-86, Stage C-106 and Stage D-76). Significance was determined by Kruskal-Wallis nonparametric one-way ANOVA with Dunn’s multiple comparisons test (**** p<0.0001, *** p<0.001, ** p<0.01, *p<0.05, n.s. nonsignificant).

We used logistic regression to model “HCC-detect” M score as a predictor of probability of HCC (Fig. 6C) and computed a predicted probability for each person using the logistic regression equation (scattered plot Fig. 6D). The HCC samples cluster around the probability of 1, few CHB samples are predicted a probability of 1 while most of the samples of the healthy and CHB samples median is around a predicted probability of 0.5. The AUC for the “HCC-spec” M score is 0.89 with specificity of 91% and sensitivity of 72% (Fig. 6D). We computed the logistic regression equation for “HCC-spec” M-scores (Fig. 6E) and the predicted probability for each person (Fig. 6F).

We then computed a combined probability score for cancer detection and HCC specification. We computed ROC curve of the combined probability of “HCC-detect” and “HCC-spec” scores of 46 healthy controls and 302 HCC patients (Fig. 7A). The calculated AUC is 0.9432 the specificity is 95% and the sensitivity is 85%. A perfect combined score is 2 which indicates a predicted probability of 1 for cancer and predicted probability of 1 that the cancer is HCC. The median score for each of the HCC stages including early stages approach 2 and is significantly different than the healthy and chronic hepatitis groups, which are not different from each other (non-parametric ANOVA and Dunnett correction for multiple comparisons) (Fig. 7B and a scatter plot for all individual samples in Fig. 7C). We calculated a threshold sum probability from the AUC curve (1.337) and used it to classify the samples as either HCC (1) or no HCC (0) (scatterplot Fig, 7D). This threshold accurately classifies 95% of the control samples 75% of the Stage A samples, 84% of the stage B samples, 82% of the stage C samples and 94% of the stage D samples (heatmap presenting the classification for each of the 398 samples (HCC-red, no HCC-blue) is presented in Fig. 7E). Using this threshold, 12% of the chronic hepatitis B (CHB) samples are classified as HCC compared with 5% of controls. The higher fraction of chronic hepatitis B that are classified as HCC compared to healthy controls might reflect the increased risk of conversion of chronic hepatitis B to HCC.

**Fig. 7.**
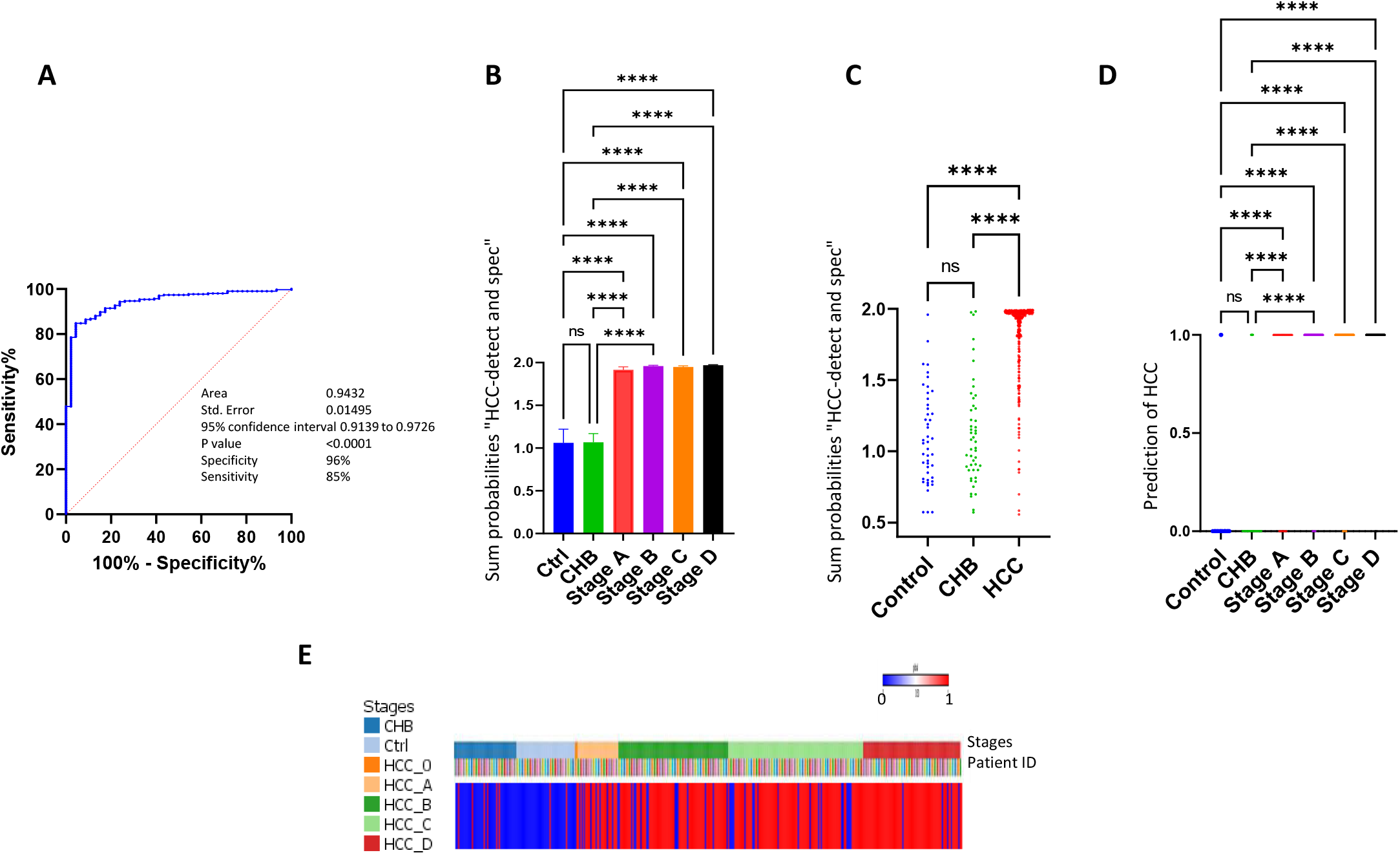
Classification of HCC by a combined “HCC-detect” and “HCC-spec” probability score (“epiLiver”). **A**. ROC curve of “HCC-detect and spec” sum probabilities score as a classifier of HCC (healthy controls, n=46 and HCC cases, n=302) in the Dhaka clinical study. **B**. Median +/- confidence intervals for the sum probabilities scores for the healthy controls, chronic hepatitis B and 4 stages of HCC. **C**. Scatter plot of the sum probabilities score for each of the samples in the control, CHB and HCC groups. **D**. Scatter plot of HCC classifications (1,0) for the samples included in the control, CHB and HCC stages groups. **E**. Heatmap presentation of HCC classifications for each of the samples in the Dhaka study. Samples were classified either as HCC (red) or nonHCC (blue). Stages are color coded as indicated in the legends. Patient IDs are color coded as indicated.

## Discussion

The main challenge for cfDNA based cancer prediction is the limited amount of tumor DNA in plasma and its dilution with DNA from blood and other tissues. Although recent technological advances cited above have boosted the feasibility of this approach, cfDNA still possess a formidable challenge particularly its translation into a widespread health management instrument which needs to be cost effective, high throughput and specific for cancer type in order to have an impact on public health. DNA methylation profiles can accurately differentiate tumor DNA from normal adjacent tissue and blood. However, tumor DNA methylation profiles might be like the pattern of methylation in unrelated tissue. Since the origin of DNA in cfDNA is unpredictable and its relative abundance is unknown, this could potentially confound the results and lead to false positives. In addition, quantitative differences in DNA methylation which are clearly sufficient to differentiate between relatively homogenous samples are confounded in plasma where mixtures of DNA from different origins with different levels of methylation coexist. Although algorithms for deriving cell of origin from DNA methylation profiles were developed, they require a relatively large number of sites which is unfeasible for a high-throughput and cost-effective public health tool. An additional complicating factor is that several cancer DNA methylation biomarkers developed to date could detect several tumors beyond HCC (Fig. 3 B, C), thus providing no direction as to the treatment response to the potential cancer detection. In order to address these challenges, we set a path to discovery that was based on the following guidelines: First, reduce cost and increase high throughput by discovering a small number of robust DNA methylation positions which are amenable to simple targeted amplification and multiplexed next generation sequencing. Second, reduce background of normal tissue DNA by focusing on sites that are ubiquitously methylation resistant across tissues and blood. Third, combine markers that detect cancer with markers that identify the tissue of origin. Markers of tissue of origin are different than cancer specific markers and do not necessarily differentiate cancer from normal cells from the same tissue of origin. Hence, the need for using two different strategies for discovery of markers that detect cancer and others that identify the tissue of origin. We demonstrate here a shortlist of methylation resistant sites that are ubiquitously unmethylated across hundreds of individual samples in 16 tissues and blood. These sites are highly enriched in CG dense promoters and depleted in enhancers and other genomic features. The depletion of methylation in certain genomic features has been recognized since the discovery of CG islands four decades ago [24] and was further confirmed by genome wide bisulfite mapping studies[25]. We show here that some of these highly DNA methylation resistant CGs are consistent across hundreds of people and thus offer a clean background. Using a training cohort from GSE54503 DNA methylation collection, we show that a fraction of these highly methylation resistant sites are methylated in HCC tumors (Fig. 1A). With main goal of cost efficiency and high throughput potential we determined the smallest number of CGs that are sufficient to serve as classifier of HCC with great accuracy. We show that methylation of only 4 CG positions is sufficient to call 98% of the HCC samples with 100% specificity. Although not all HCC samples have all 4 positions methylated, 98% of the samples have at least one CG methylated. We proposed that these 4 CGs serve as “HCC-detect” set of markers. We validated that these markers could classify HCC accurately with high sensitivity and specificity using a dataset of 739 HCC and samples from 2212 healthy tissues. However, as expected when “HCC-detect” was used against a panel of 31, cancers from other tissue origins were detected at different sensitivities (Fig.2A). To provide a classifier of liver origin tumor we discovered a single DNA methylation biomarker that distinguished DNA from liver origin from other tissues which we termed “HCC-spec”. A combined test of “HCC-spec” and “HCC-detect” provides a highly accurate classification of HCC and distinguishing it from other tumors providing clear guidance on the location of the tumor, which will speed up applying the necessary follow-up and clinical measures. Importantly, both “HCC-detect” and “HCC-spec” markers are methylated at the early stages of HCC and remain methylated to the most advanced stages. This will provide a broad-spectrum single test for first screening of asymptomatic patients notwithstanding the stage of cancer avoiding missing patients who have moved to a later stage or who are at an earlier stage of the disease. “HCC-detect” and “HCC-spec” markers were discovered and validated in tumor biopsy material, it is important to determine whether they are methylated in plasma cfDNA as well. We therefore validated the methylation state of these markers in a previously published data set that examined genome wide methylation of HCC in tumor and plasma (Fig. 4A-E) [23]. These data are consistent with other studies that showed correspondence between DNA methylation profiles of tumor tissue and plasma cfDNA [17, 19].

We applied the agnostic criteria of ubiquitous methylation resistance in normal tissues and methylation in HCC for selecting the “HCC-detect” biomarker set, irrespective of biological function. Nevertheless, the fact that we found a set of CGs that are methylated in a large subset of HCC patients and at all stages of HCC suggests that these DNA methylation events might be important for the early stages of the disease as well as its maintenance. The fact that no single marker is methylated in all normal samples but at least one of the 4 markers is methylated in almost all HCC patients suggests that at least one of the 4 markers is essential. Indeed, the “HCC-detect” CGs are associated with genes involved in cell cycle regulatory events, EMT transition and angiogenesis. *CHFR* encodes a cell cycle (G2/M) checkpoint, which was suggested to be a tumor suppressor gene[26]; methylation of *CHFR* was associated with non-invasive colorectal cancer[27], esophageal carcinoma[28], hepatocellular carcinoma[29], higher grade gastric cancer[30] and non-small cell lung carcinoma[31]. *Vasohibin 2 (VASH2*) gene was implicated in angiogenesis in invasive tumors [32] and was previously shown to be methylated in HCC[33]. *Vash2* is a promoter of an angiogenesis gene it is therefore expected to promote cancer and indeed higher expression of *Vash2* is associated with poor progression of pancreatic cancer[34], esophageal squamous cell carcinoma[35], breast cancer[36] and epithelial-mesenchymal transition (EMT) in HCC[37]. We would therefore expect hypomethylation of the 5’regulatory region of the gene rather than the observed hypermethylation of the 1^st^ exon. We don’t know how this methylation event is correlated with expression and it needs to be further explored. *Cyclin-J gene* (*CCNJ*) encodes the cyclin J protein which was proposed to be involved in early embryonic division cycles of Drosophila[38] and was previously found to be methylated in liver cancer[39]. *Glutamate Receptor, Ionotropic, Delta 2* (*Grid2*) *Interacting Protein 1 gene* (*GRID2IP*) is expressed in the brain it interacts with Glutamate receptor delta-2 and was shown to be involved in synaptogenesis in fiber-Purkinje cells[40]. However, its role in cancer is not yet fully explored. The methylated CG included in our detect marker set in in a CG island in the body of the gene and it is unclear whether this CG island has any regulatory role. The role of this gene in HCC remains to be determined. The *F12* gene encodes coagulation factor XII[41]. The CG position in a CG rich island in the body of this gene and was selected here only for its exquisite HCC specific methylation and methylation resistance in 31 other tumors (Fig. 2D). The relationship between methylation and expression is unclear and needs to be further explored.

We then examined whether these data could be applied as a high throughput cost effective assay to detect HCC in a clinical setting by examining plasma DNA from 402 people which included all stages of HCC as well as healthy controls and chronic hepatitis B patients that are at high risk of conversion to HCC. By targeting a small number of amplicons, we significantly reduce the cost and increase the number of potential reads per sample and our assay is enabled for high throughput and automated formats. We had only 4 cases out of the 402 that didn’t provide sufficient reads (a threshold of 100 reads per gene) in at least one of genes. The average coverage per sample for each of the gene regions ranged from 3160 to 5181, suggesting that most plasma samples from either healthy controls or cases have sufficient DNA to generate methylation information on these genes. All 5 CG positions included in “HCC-detect” and “HCC-spec” are significantly more methylated than in either healthy controls, chronic hepatitis B or both (CCNJ is nominally significant when compared to healthy controls and significant against chronic hepatitis B). As expected, most control samples are completely unmethylated with a median methylation of less than 0.5%. Methylation in HCC is heterogenous ranging from baseline levels to 80% (Fig. 5 A). This probably reflects to some extent random heterogeneous mixture of tumor and normal cfDNA in plasma but also the heterogeneity of methylation of these sites in the original tumor (Fig. 1 and 2).

Our markers are CG rich and by sequencing a 100-200 bp region proximal to the CG IDs that were discovered in the Illumina 450K array data we capture information on the state of methylation of several neighboring CGs (Fig. S1B). Our data indicates that in HCC samples the levels of methylation across CGs in each of the five sequenced regions are correlated while only sporadic methylation is detected in the healthy controls which might be biological or just a result of spurious infrequent errors in bisulfite conversion or sequencing. By examining the entire profile, we can increase our specificity and exclude such cases. To reflect the consistency of methylation across the region we calculated the Median for each CG (as is visualized in heatmap in Fig. 5B) The medians for each gene region (Fig. 5B) and the “HCC-detect” sum (Fig. 5B) clearly separate the HCC samples from chronic hepatitis B and healthy controls (Fig. 5B). Since our training set analysis suggested that methylation in any one of the 4 CGs is sufficient to classify a sample as HCC we have given equal weight to each region and used the normalized sum of the medians as the “HCC-detect” M score and the normalized median for *F12* region as the “HCC-spec” M score. Logistic regression of M scores calculated (Fig. 6B, E) the predicted probability of HCC for each sample; 89% of the HCC samples scored close to 1 at all stages in contrast to 4% of the healthy controls which were misclassified as HCC (Fig. 6C). A combined “HCC-detect” and “HCC-spec” score which accurately classified predicted probability had an ROC of 0.9432 (Fig. 7A), (Fig. 2D, E) classified HCC from other cancers in the TCGA data, classified 256 out of the 303 HCC samples as HCC (84.5%) and misclassified 2 out of 46 healthy controls as HCC (Fig. 7 C,D,E). Our results validated the “HCC-detect” and “HCC-spec” markers, in cfDNA clinical samples and provided a way forward for developing a feasible, high-throughput and cost effective test for noninvasive screening and detection of HCC that is tumor specific. We termed the combined “HCC-detect” and “HCC-spec” “epiLiver”.

The sensitivity of the CF plasma DNA test was lower than in the TCGA data 85%-89% in the clinical study compared to 96 to 98% in the analysis of TCGA DNA methylation values. There are a few possible reasons. First, TCGA data is derived from tumor tissue while cfDNA that originates from tumors is mixed with DNA from blood and other normal tissues in unpredictable ratios. Second, the amount of cfDNA varies across patients. Third, quality of plasma derived in clinical settings is probably not even and different levels of mixture of genomic DNA might be caused by different handling of samples. Fourth, different genomic regions might be heterogeneously represented in cfDNA in different samples. Nevertheless, our assay demonstrates high sensitivity and specificity as well as cost effectiveness and high throughput. One of the challenges of early noninvasive tumor detection is defining the tissue of origin. Remarkably only one CG was sufficient to accurately reveal the tissue of origin suggesting that a cost-effective screen for specific tumor types might be feasible. However, full assessment of the clinical value of the “HCC-detect” and “HCC-spec” markers for early detection of HCC requires an adequately powered prospective study.

Although the 5 regions included in the “HCC-spec” and “HCC-detect” are devoid of methylation in the vast majority of healthy controls as is the case in the TCGA data, a small number of healthy normals (2 in the combined “HCC-detect” and “HCC-spec” score) had a profile that resembled HCC patients with consistent methylation across all CGs. At this stage we don’t know whether these false positives are truly “false” or whether they represent undetected cases. Our clinical study did not include follow up of such cases. One of the challenges of early detection is to further study and understand these “false positives” as well as deciphering the exact boundary between healthy and controls, lowering the threshold will increase the sensitivity of detection but this should be done without increasing the level of “false discoveries”. Although the vast majority of hepatitis B patients had very low level of methylation and there is no statistically significant difference noted between healthy controls and CHB patients mean M score, there was a higher fraction of HCC classifications in the chronic hepatitis B group (12%). The fact that we got a higher fraction of HCC calls in the HepB group might be consistent with the higher risk of conversion to HepB; follow up of these cases is warranted.

In summary, our study reveals a feasible, cost effective and accurate noninvasive test for detection of HCC. Applying this test for screening people at risk for developing HCC could potentially have an important impact on relieving the burden of this tumor on the individual as well as the health system.

## Data Availability

All data will be available upon request.

## Acknowledgment

The results shown here are in part based upon data generated by the TCGA Research Network: https://www.cancer.gov/tcga.

## Materials and Methods

### DNA methylation public data

Normalized DNA methylation beta values for 450K sites included in Illumina DNA methylation arrays for a total of 11,636 samples from healthy blood, normal tissues, cancer tissues and noncancer associated tissues NAT were downloaded from TGCA and GEO sites as listed in table 1. DNA methylation differences at specific CG positions between the groups were computed using t statistics adjusted by Bonferroni corrections for 450K multiple tests.

### Clinical study design

Study protocol was approved by IRB board of icddr,b (Dhaka, Bangladesh) study protocol PR-18025. 402 participants were recruited from the Dhaka area to the study included 49 healthy controls, 51 Chronic hepatitis B patients and 303 HCC patients from stages 0 to D (HCC O n=2, HCCA n=32, HCC B n=86, HCC C n=106, HCC D n=77 (See Table 2 for demographics). The age of the healthy control and the chronic Hepatitis B (CHB) groups were significantly younger than the HCC group, however there was no significant effect of age on HCC prediction by DNA methylation as determined by a logistic regression in either the HCC or control groups. However, in the CHB group age had a significant effect on HCC prediction by the DNA methylation test, consistent with the idea of a higher probability for older patients with chronic hepatitis to convert to HCC. There were no differences between the groups in the sex distribution. However, there was a somewhat significant lower alcohol use and higher fraction of smokers in the HCC groups (Table 2). HCC staging was diagnosed according to EASL–EORTC Clinical Practice Guidelines: Management of hepatocellular carcinoma [42]. Hepatitis B diagnosis was confirmed using AASLD practice guideline for chronic Hepatitis B (https://www.aasld.org/publications/practice-guidelines). All participants were properly informed about the study and have signed the informed consent form approved by the icddr,b IRB. Inclusions criteria were participants of either sex 18 to 70 years of age, confirmed diagnosis of HCC using EASL-EORTC guidelines and chronic hepatitis B using AASLD guidelines, non-metastatic liver cancer, Hepatitis B surface antigen positive by ELISA and persistence of > 6 months. Exclusion criteria were unwilling or unable to provide informed consent, unwilling or unable to comply with requirements of protocol, participation in a different clinical trial investigating a vaccine, drug, medical device or medicinal procedure less than 4 weeks preceding the current study, planned participation in another clinical trial during present study period, known case of cirrhosis, any other known inflammatory disease (bacterial or viral infection with the exception of hepatitis B or C), known case of diabetes, asthma, autoimmune disease, any other diagnosed cancer, for healthy controls any known inflammatory or infectious disease including Hepatitis B and Hepatitis C and any diagnosis of chronic disease, cancer medication use or drugs of abuse. Patients were assigned an ID that was kept confidential according to hospital regulations and identity was revealed only to approved hospital personnel. Participants provided consent for DNA methylation biomarker research. Blood sample collection ad plasma separation was performed at icddr,b in Dhaka Bangladesh and was then shipped to HKG epitherapeutics for further analysis. The HKG epitherapeutics lab team was blinded on the identity of the samples throughout the lab analytic procedures. Data was then analyzed in Montreal and shared with icddr,b who provided the results to the respective clinical personnel.

### Preparation of CF plasma DNA

Blood was collected in 9-ml tubes containing K3-EDTA and processed within 1 h. Plasma and peripheral blood monocyte separation was performed according to GE Healthcare Cat No 71=7167-00 protocol. Plasma was frozen and shipped. Plasma samples (1 ml) were thawed, and DNA was extracted by previously described guanidine isothiocyanate method[43] and binding to silica magnetic beads followed by 80% ethanol washes and water elution.

### Multiplexed Targeted DNA methylation Illumina amplicon sequencing

Bisulfate conversion was performed using EZ-96 DNA Methylation MagPrep (D5041, Zymo Research) followed by two rounds of polymerase chain reaction. For the first round we used primers that included an anchoring sequence and sequences targeting cg02012576 (*CHFR*), cg03768777 (*VASH2*), cg05739190 (*CCNJ*), cg24804544 (*GRID2IP*) and cg14126493 (*F12*) using Bio-Rad C1000 Touch Thermal Cycler (Bio-Rad Laboratories, CA, USA) (the primers are available upon request). 5 microliters of the first PCR reaction were subjected to a second round of PCR amplification using primers containing indexes for barcoding the samples (the primers are available upon request). PCR products were pooled, and the pooled library was then purified twice using AMPure XP Beads (Beckman Coulter Life Sciences, CA, USA) and quantified by RealTime PCR using NEBNext® Library Quant Kit for Illumina (New England Biolabs, MA, USA). Sequencing was performed on the Illumina platform using MiSeq Reagent Nano Kit V2 (Illumina, CA, USA).

### Statistical methods

W**e** used the computing environment R version 3.4.4. For penalized regressions we used the R statistical package “penalized” [44], for multivariable linear regression analysis we used the lm function in R to fit linear models and for genomic feature enrichment we performed a hypergeometric test using phyper function in R. For other statistical analyses we used Graph Pad Prism 9.01 statistical package. Normality and log normality were tested using Shapiro-Wilk and Kolmogorov-Smirnov tests. Nonparametric statistics were used to test significance when data failed normality tests. For two group comparisons a two tailed Mann Whitney test was used and for multiple group comparisons we used Kruskal-Wallis test followed by Dunn’s multiple comparisons test to derive the adjusted p value. ROC was computed using the ROC test in GraphPad. To generate heatmaps we used GENE E software from the Broad institute (https://software.broadinstitute.org/GENE-E/).

## Figure legends

**Fig. S1.**
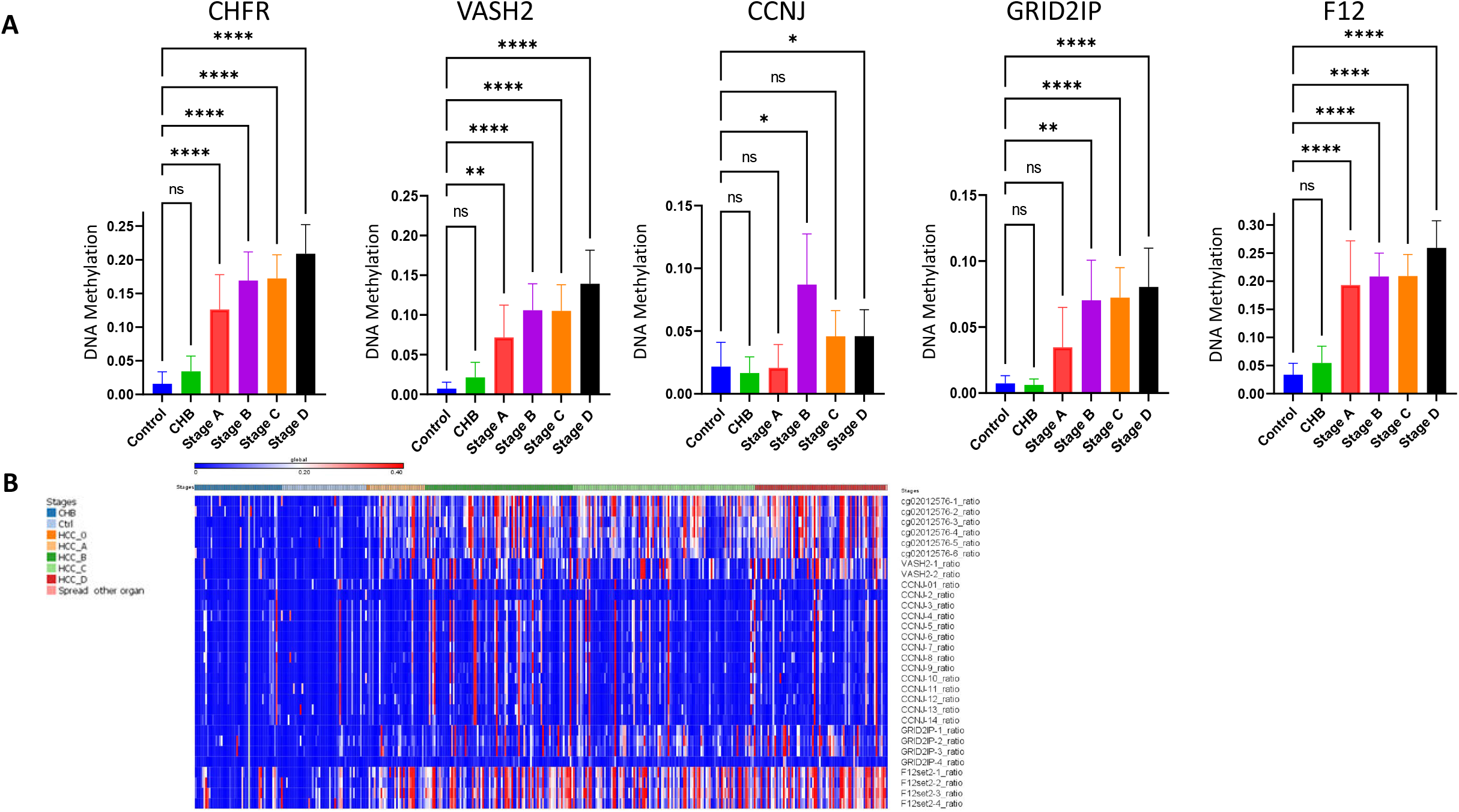
Differential methylation of “HCC-detect” and “HCC-spec” CGs at different stages of HCC in the Dhaka clinical study. Mean methylation and confidence intervals for the 4 CGs included in the “HCC-detect” set (cg02012576 (CHFR), cg03768777 (VASH2), cg05739190 (CCNJ), cg24804544 (GRID2IP) and “HCC-spec” cg14126493 (F12) (n for each group as indicated in Fig. 5). B. heat map depicting the methylation values for all CGs that were included in the sequenced regions for all 5 genes.

## Supplemental tables

**Table S1.**
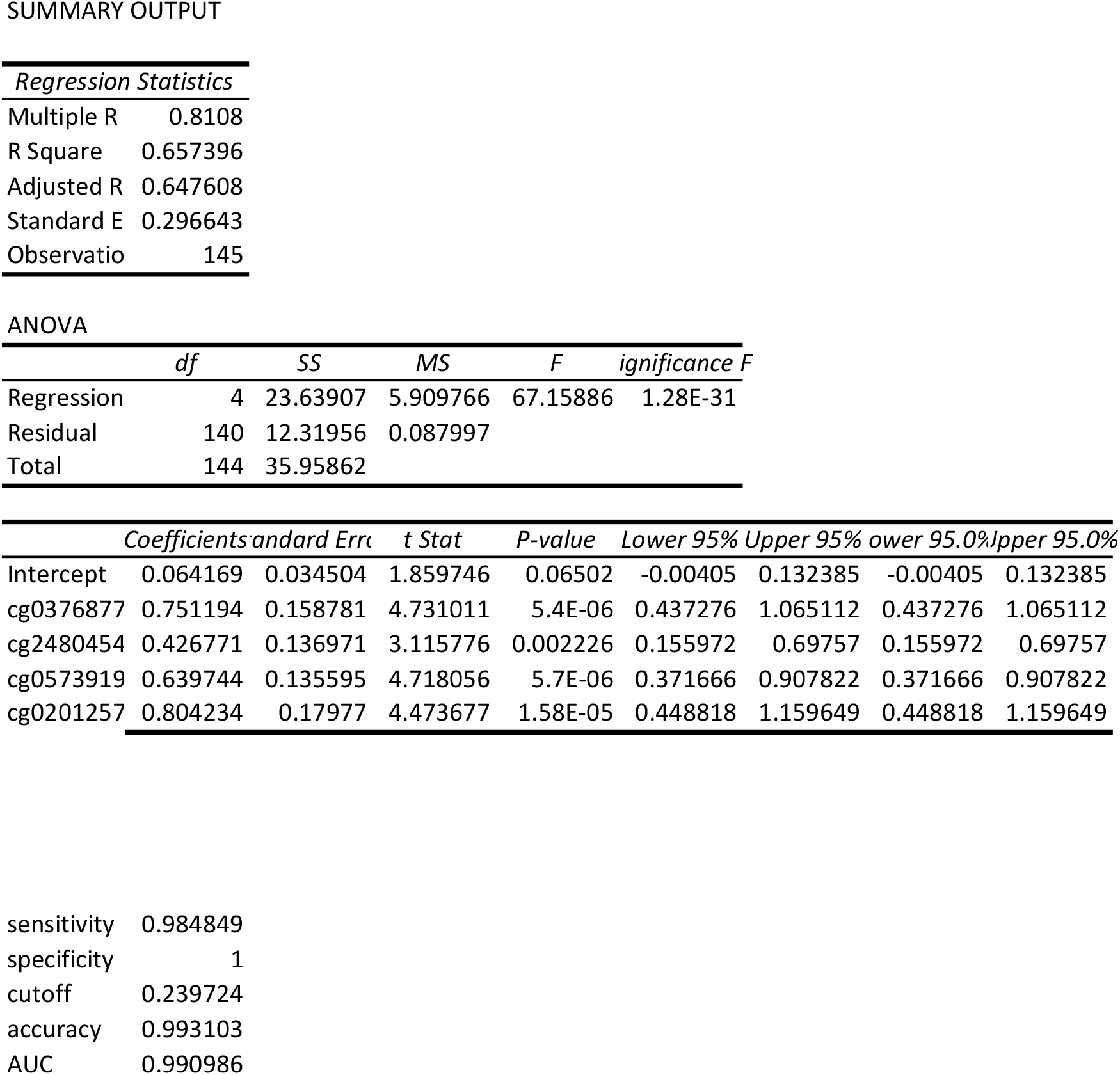
Model statistics for “HCC-detect”.

SUMMARY OUTPUT
| *Regression Statistics* |  |
| --- | --- |
| Multiple R | 0.8108 |
| R Square | 0.657396 |
| Adjusted R | 0.647608 |
| Standard E | 0.296643 |
| Observatio | 145 |

ANOVA
|  | *df* | *SS* | *MS* | *F* | *ignificance F* |
| --- | --- | --- | --- | --- | --- |
| Regression | 4 | 23.63907 | 5.909766 | 67.15886 | 1.28E-31 |
| Residual | 140 | 12.31956 | 0.087997 |  |  |
| Total | 144 | 35.95862 |  |  |  |

|  | *Coefficients* | *andard Error* | *t Stat* | *P-value* | *Lower 95%* | *Upper 95%* | *ower 95.0%* | *pper 95.0%* |
| --- | --- | --- | --- | --- | --- | --- | --- | --- |
| Intercept | 0.064169 | 0.034504 | 1.859746 | 0.06502 | -0.00405 | 0.132385 | -0.00405 | 0.132385 |
| cg0376877 | 0.751194 | 0.158781 | 4.731011 | 5.4E-06 | 0.437276 | 1.065112 | 0.437276 | 1.065112 |
| cg2480454 | 0.426771 | 0.136971 | 3.115776 | 0.002226 | 0.155972 | 0.69757 | 0.155972 | 0.69757 |
| cg0573919 | 0.639744 | 0.135595 | 4.718056 | 5.7E-06 | 0.371666 | 0.907822 | 0.371666 | 0.907822 |
| cg0201257 | 0.804234 | 0.17977 | 4.473677 | 1.58E-05 | 0.448818 | 1.159649 | 0.448818 | 1.159649 |
| sensitivity | 0.984849 |
| specificity | 1 |
| cutoff | 0.239724 |
| accuracy | 0.993103 |
| AUC | 0.990986 |

**Table S2.** List number of samples and acronyms of other cancers analyzed.

| **TCGA** | Study Abbreviation | Study Name | Cancer Subject number | NAT subject r | Total |
| --- | --- | --- | --- | --- | --- |
| **TCGA-ACC** | ACC | Adrenocortical carcinoma | **80** | 0 | 80 |
| **TCGA-BLCA** | BLCA | Bladder Urothelial Carcinoma | **412** | 21 | 433 |
| **TCGA-BRCA** | BRCA | Breast invasive carcinoma | **796** | 95 | 891 |
| **TCGA-CESC** | CESC | Cervical squamous cell carcinoma and endocervical adenocarcinoma | **309** | 3 | 312 |
| **TCGA-CHOL** | CHOL | Cholangiocarcinoma | **36** | 9 | 45 |
| **TCGA-COAD** | COAD | Colon adenocarcinoma | **315** | 38 | 353 |
| **TCGA-DLBC** | DLBC | Lymphoid Neoplasm Diffuse Large B-cell Lymphoma | **48** | 0 | 48 |
| **TCGA-ESCA** | ESCA | Esophageal carcinoma | **186** | 16 | 202 |
| **TCGA-GBM** | GBM | Glioblastoma multiforme | **153** | 2 | 155 |
| **TCGA-HNSC** | HNSC | Head and Neck squamous cell carcinoma | **530** | 50 | 580 |
| **TCGA-KICH** | KICH | Kidney Chromophobe | **66** | 0 | 66 |
| **TCGA-KIRC** | KIRC | Kidney renal clear cell carcinoma | **325** | 159 | 484 |
| **TCGA-KIRP** | KIRP | Kidney renal papillary cell carcinoma | **276** | 45 | 321 |
| **TCGA-LAML** | LAML | Acute Myeloid Leukemia | **140** | 0 | 140 |
| **TCGA-LGG** | LGG | Brain Lower Grade Glioma | **534** | 0 | 534 |
| **TCGA-LIHC** | LIHC | Liver hepatocellular carcinoma | **380** | 50 | 430 |
| **TCGA-LUAD** | LUAD | Lung adenocarcinoma | **411** | 22 | 433 |
| **TCGA-LUSC** | LUSC | Lung squamous cell carcinoma | **337** | 36 | 373 |
| **TCGA-MESO** | MESO | Mesothelioma | **87** | 0 | 87 |
| **TCGA-OV** | OV | Ovarian serous cystadenocarcinoma | **10** | 0 | 10 |
| **TCGA-PAAD** | PAAD | Pancreatic adenocarcinoma | **185** | 10 | 195 |
| **TCGA-PCPG** | PCPG | Pheochromocytoma and Paraganglioma | **184** | 3 | 187 |
| **TCGA-PRAD** | PRAD | Prostate adenocarcinoma | **503** | 50 | 553 |
| **TCGA-READ** | READ | Rectum adenocarcinoma | **99** | 7 | 106 |
| **TCGA-SARC** | SARC | Sarcoma | **265** | 4 | 269 |
| **TCGA-SKCM** | SKCM | Skin Cutaneous Melanoma | **473** | 2 | 475 |
| **TCGA-STAD** | STAD | Stomach adenocarcinoma | **395** | 2 | 397 |
| **TCGA-TGCT** | TGCT | Testicular Germ Cell Tumors | **156** | 0 | 156 |
| **TCGA-THCA** | THCA | Thyroid carcinoma | **515** | 56 | 571 |
| **TCGA-THYM** | THYM | Thymoma | **124** | 2 | 126 |
| **TCGA-UCEC** | UCEC | Uterine Corpus Endometrial Carcinoma | **439** | 45 | 484 |
| **TCGA-UCS** | UCS | Uterine Carcinosarcoma | **57** | 0 | 57 |
| **TCGA-UVM** | UVM | Uveal Melanoma | **80** | 0 | 80 |
| **Total** |  |  | 8906 | 727 | 9633 |

**Table S3.** Model statistics for “HCC-spec”.

SUMMARY OUTPUT
| *Regression Statistics* |  |
| --- | --- |
| Multiple R | 0.798739 |
| R Square | 0.637984 |
| Adjusted R | 0.633674 |
| Standard E | 0.257195 |
| Observatio | 86 |

ANOVA
|  | *df* | *SS* | *MS* | *F* | *ignificance F* |
| --- | --- | --- | --- | --- | --- |
| Regression | 1 | 9.792313 | 9.792313 | 148.034 | 3.16E-20 |
| Residual | 84 | 5.556524 | 0.066149 |  |  |
| Total | 85 | 15.34884 |  |  |  |

|  | *Coefficients* | *andard Error* | *t Stat* | *P-value* | *Lower 95%* | *Upper 95%* | *ower 95.0%* | *pper 95.0%* |
| --- | --- | --- | --- | --- | --- | --- | --- | --- |
| Intercept | 0.179496 | 0.055716 | 3.221603 | 0.001814 | 0.068698 | 0.290294 | 0.068698 | 0.290294 |
| cg1412649 | 1.346715 | 0.110687 | 12.16692 | 3.16E-20 | 1.126602 | 1.566827 | 1.126602 | 1.566827 |

**Table S4.**
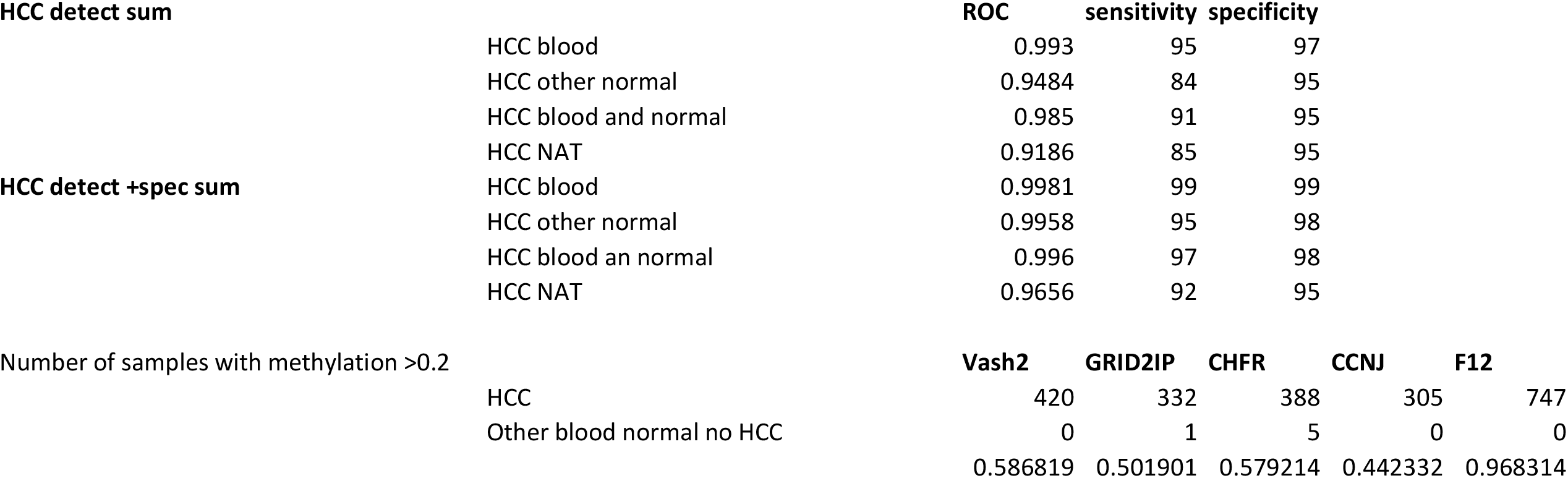
ROCs for “HCC-detect” sum “HCC-spec” + “HCC-detect” sum scores and number of samples with methylation values larger than 0.2.

## Footnotes

1 **www.cancer.org/cancer/livercancer**.

